# Evaluation of implementing a school-based vision care program using mobile eye exam lanes

**DOI:** 10.64898/2025.12.25.25343010

**Authors:** Jacob M Perkins, Brian Grover, Diamond R. Guy, Mohammed Mehdi Shahid, Talia Gearinger, Matthew Gearinger, Brittany Wong, Rajeev S. Ramchandran

## Abstract

**Purpose:** Visual impairment in children is a significant public health issue, especially for those from disadvantaged backgrounds. Vision screening by schools and pediatricians have had limited success in resolving this concern. This study examined the outcomes, challenges, and demographics of the MobilEyes program, a school-based mobile eye care initiative in Rochester, New York.

**Methods:** During the 2022-2023 academic year, students in PreK, K, 1st, 3rd, 5th, and 7th grade underwent visual acuity screening per New York State Guidelines. A positive screen was visual acuity <20/40 for kindergarteners or younger and <20/30 for 1st graders or older. Children who failed vision screening and whose parents provided consent for school-based eye examination subsequently underwent cycloplegic refraction and dilated fundus examination by a pediatric ophthalmologist. Demographics subgroups were analyzed by school, school region, and grade level.

**Results:** 1399 students from six schools underwent vision screening, of which 387 (27.7%) failed the initial screening. From those, 125 (32.3%) returned signed parental consent forms and 108 (27.9%) underwent full eye examinations. Of this group, 68 (63.0%) were prescribed glasses, and 40 (37.0%) did not require glasses prescription and were considered false positives. Significantly more urban students failed their vision screening than suburban students, 32.8% vs 22.5% (p<0.001), and urban students had a lower false positive vision screening result (meaning that they failed their vision screening but were found to have a normal exam) than suburban students 27.6% vs 48% (p=0.047). Across all schools we found statistically significant differences between grade levels in the number failing vision screening (p<0.001) and returning consent forms (p=0.021).

**Conclusion:** Our data highlights the need for additional support of school screening and full eye examinations, especially in urban regions. Future studies should address the challenges of high vision screening false positivity percentages and obtaining parental consents to perform eye examinations.

## Introduction

Visual impairment in children remains a major public health challenge worldwide. Globally over 200 million children have refractive error and in the United States roughly 3% of children younger than 18 years old experience visual impairment or blindness (1–4). Recent analysis of the National Survey of Children’s Health shows that the prevalence of pediatric vision screening has significantly declined in the U.S. by 9.5% (69.6% to 60.1%, *P* < 0.001) across 5-years (2016–2020). According to the same report, children are also receiving less ophthalmic care (4). Furthermore, children from disadvantaged backgrounds are disproportionately impacted (2, 4–6). Minoritized children and those living below the 400% Federal Poverty Level receive fewer and less intensive eye care services, as indicated by lower expenditures (7). One study showed that students from Title I schools (which receive federal financial assistance for children from low-income families) failed one subset of vision screening tests at 2.4 and 1.9 times the rate of control students with two different screening methods (8). These findings raise concern given the worse academic and quality life outcomes associated with uncorrected vision problems (9, 10).

Although 41 states mandate at least one community or school-based pediatric vision screening, only six states require a vision screening annually or biannually. Across states, there are significant variations in screening methods and criteria (80% test distance visual acuity, 22% test color vision, 20% test near visual acuity), screening location (22% in the community setting, 59% in schools, 20% in both) and the grade levels screened (80% in kindergarten, 59% in middle school, 37% in high school). This lack of standardization and wide variation in state regulations points to a need for the development of evidence-based criteria for vision screening programs for school-aged children (2).

To address these deficits, efforts to develop and improve pediatric vision screening programs are underway across the United States. Children that do not have a consistent source of health care are associated with 46% lower odds of vision testing. In Alabama, of the children 3-18 years old on Medicaid who attended a well care check, 42% also had a vision screening with >98% occurring at the same visit (11). Unfortunately, at pediatric primary care facilities it is estimated that children miss 30% to 50% of their well child visits. Furthermore, minoritized populations that are socioeconomically disadvantaged and miss a greater proportion of these visits (12). School-based vision screening programs aim to increase the identification of children with vision problems and some are performed in conjunction with school-based eye exams to increase access to pediatric eyecare. These “mobile eye care” programs are supported by an eye care team outfitted with portable eye exam equipment (5, 10, 13–16).

MobilEyes is a population health focused approach to promoting and achieving eye health across the life span, started by the Flaum Eye Institute at the University of Rochester, Rochester, NY, USA. Its pediatric focus is on school-based vision screenings and exams for students in underserved communities from low income, minoritized, or rural backgrounds. MobilEyes performs screenings at schools in the Greater Rochester, NY region which serve a diverse student population (Black: 18-74%, Hispanic: 21-62%, White: 2-46%) where 58-97% of students are economically disadvantaged (17). In the current analysis, we aim to evaluate the outcomes, challenges, and demographics of MobilEyes for the first full school year with full in person classes in New York State since the COVID-19 pandemic was deemed over by the Centers for Disease Control. The findings from relaunching this school based vision screening and eye exam program will be compared with similar programs in the United States to highlight challenges and opportunities of such programs.

## Materials and Methods

Throughout the 2022-2023 school year, the MobilEyes program visited six different schools in Rochester, NY. Students in grades PreK, K, 1st, 3rd, 5th, and 7^th^ underwent visual acuity screening per New York State guidelines. Children in other grades were seen based on teacher concern for those grades or if the students were new to the school. All children present at school on the day(s) of screening underwent evaluation on a class-by-class basis. Screening was performed by a team consisting of a pediatric ophthalmologist, medical students, physician assistant students, and/or ophthalmic technicians. Testing included monocular distance visual acuity and binocular near visual acuity measurement via Snellen chart or Allen figures per New York State vision screening guidelines. Failed vision screening test was defined as visual acuity in either eye poorer than 20/40 for preschoolers and kindergarteners, or poorer than 20/30 for first graders and higher. Children who did not pass the screening were sent home with a consent form and information packet regarding upcoming full eye examinations. Color vision was also tested as it is mandated by New York State, but an abnormal color vision test alone was not considered a vision screening failure and follow-up eye care was not recommended for these cases.

Children with signed consent from parents/guardians were eligible to undergo a full eye examination by a pediatric ophthalmologist that was billed to the health insurer covering the child. Parents/guardians were notified that dilated eye exams would be charged to the health insurance carrier covering the child and that glasses, if needed, would be provided at no additional cost to the family. Full eye examination included cycloplegic refraction. If indicated, strabismus examination was also performed. Anterior segment exam was performed either by pen light, portable slip lamp, or tabletop slit lamp. Dilated fundus examination was performed by indirect ophthalmoscopy. Based on exam findings, appropriate follow-up appointments were scheduled. Those students who were found to have significant refractive error were prescribed glasses. Based on these prescriptions, glasses were fit and delivered to the school by an optician through philanthropic funds at no cost to the family. <u>This study is a secondary analysis of the pre-existing, de-identified data gathered during this 2022-2023 academic year of the MobilEyes program and accessed on May 10^th^ 2024. This ensures that authors could not identify individual participants during or after data collection. The Research Subjects Review Board (RSRB), which is the University of Rochester’s Institutional Review Board (IRB), determined that this research was not human subject research. Therefore the study did not require RSRB review or approval, or consent to access the deidentified data.</u>

## Statistical Analysis

Pearson’s chi-squared (χ2) tests of independence were utilized to determine the relationship between categorical screening outcomes and student demographics such as grade level, school, and school region. All statistical analyses were performed using RStudio version 2023.06.0+421 (Posit, Boston, MA, USA, 2023). A value of P <0.05 was considered statistically significant.

## Results

Among 2166 eligible children in six schools and nine grades (Pre-K to 9^th^ grade), 1399 (64.5%) were present at school on the days vision screening took place by either the school nurse or the MobilEyes team, and underwent vision screening. Of the students screened, 387 (27.7%) failed one or more tests for visual acuity. For children who failed the initial screening, 125 (32.3%) parental consent forms were returned, and these students were eligible for an in-school comprehensive eye examination by a pediatric ophthalmologist. Ultimately, 108 (27.9%) participated in a full eye examination. Of the students examined, 68 (63.0%) were prescribed and provided with glasses. We identified 40 (37.0%) children whose eye exams were normal despite failing the initial visual acuity screening. These cases were considered false positives.

Across the six schools, failed screening percentage ranged from 17.1% to 38.6%. On Pearson’s chi-squared (χ2) tests of independence this difference between schools was significant (p<0.001). However, the difference in consent (29.1% to 37.0% (p = 0.916)) and false positives (23.3% to 58.3% (p = 0.184)) between schools were not statistically significant (**Table 1**).

**Table 1.** MobilEyes vision screening results across six schools. * p < 0.05, ** p < 0.01, *** p < 0.001

| School | Vision Screening (%) |  |  |  | Consent Process (%) |  |  |  | Glasses Prescription (%) |  |  |  |
| --- | --- | --- | --- | --- | --- | --- | --- | --- | --- | --- | --- | --- |
|  | Failed Screening | Passed Screening | Total | p-value | No Consent | Consent | Total | p-value | Glasses | No Glasses | Total | p-value |
| School 1 | 54 (33.0) | 110 (67.1) | 164 (11.7) | <0.001*** | 34 (63.0) | 20 (37.0) | 54 (14.0) | 0.916 | 15 (75.0) | 5 (25.0) | 20 (18.5) | 0.184 |
| School 2 | 127 (30.9) | 284 (69.1) | 411 (29.4) |  | 90 (70.9) | 37 (29.1) | 127 (32.8) |  | 23 (76.7) | 7 (23.3) | 30 (27.8) |  |
| School 3 | 49 (38.6) | 78 (61.4) | 127 (9.1) |  | 32 (65.3) | 17 (34.7) | 49 (12.7) |  | 4 (50.0) | 4 (50.0) | 8 (7.4) |  |
| School 4 | 49 (24.0) | 155 (76.0) | 204 (14.6) |  | 34 (69.4) | 15 (30.6) | 49 (12.7) |  | 6 (54.5) | 5 (45.5) | 11 (10.2) |  |
| School 5 | 29 (17.1) | 141 (82.9) | 170 (12.2) |  | 20 (69.0) | 9 (31.0) | 29 (7.5) |  | 5 (41.7) | 7 (58.3) | 12 (11.1) |  |
| School 6 | 79 (24.5) | 244 (75.5) | 323 (23.1) |  | 52 (65.8) | 27 (34.2) | 79 (20.4) |  | 15 (55.6) | 12 (44.4) | 27 (25.0) |  |
| Total N | 387 (27.7) | 1012 (72.3) | 1399 (100) |  | 262 (67.7) | 125 (32.3) | 387 (100) |  | 68 (63.0) | 40 (37.0) | 108 (100) |  |

In the school region analysis, a significantly higher number of urban students (32.8%) failed their vision screening compared to those in suburban schools (22.5%) (p<0.001). The percent of consents obtained for a full eye exam from parents of students who failed their vision screening did not vary significantly between urban (32.2%) and suburban school students (32.5%) while the percentage of false positives did (27.6% vs 48.0% (p = 0.047)) (**S1 Table**).

Controlling for potential bias we completed the same school region analysis but removed pre-K, 4th, 6th, and 7^th^ grades which were only from one school each (school 5, 2, 3, and 1 respectively) and therefore with only one region for each of those grades. Results were similar with significantly higher number of urban students (32.5%) failing their vision screening compared to those in suburban schools (24.1%) (p=0.002). There was no difference in percent of parental consents completed for an exam between urban (34.3%) and suburban school students (34.2%), but there was a significant difference in the percentage of false positive screening in urban versus suburban schools (25.0% vs 49.0% (p = 0.025)) (**Table 2**).

**Table 2.**
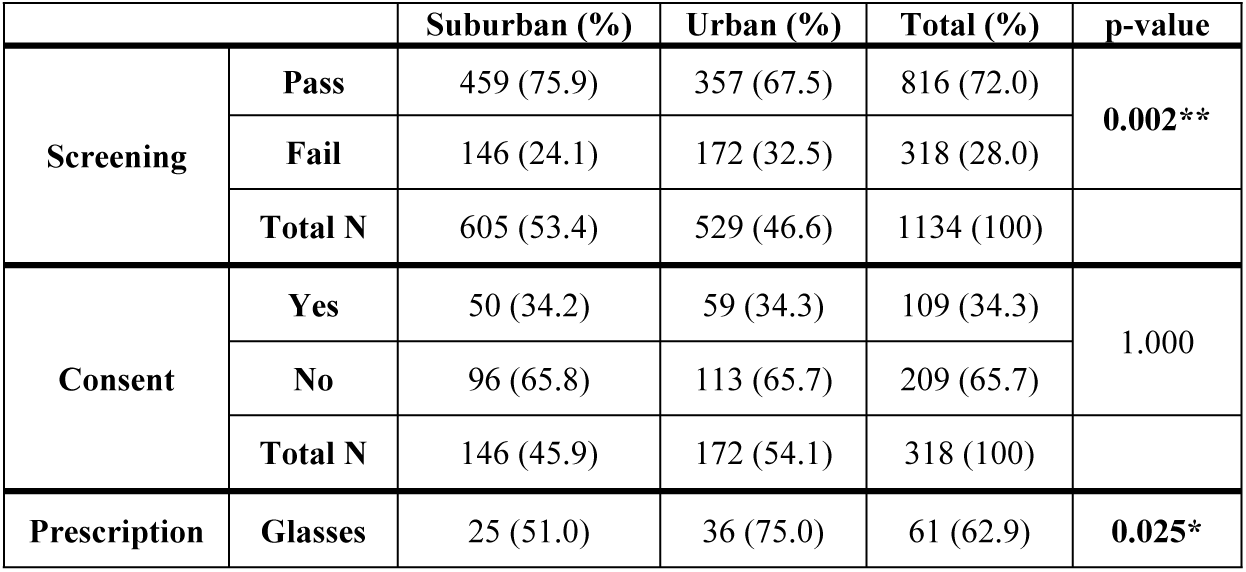
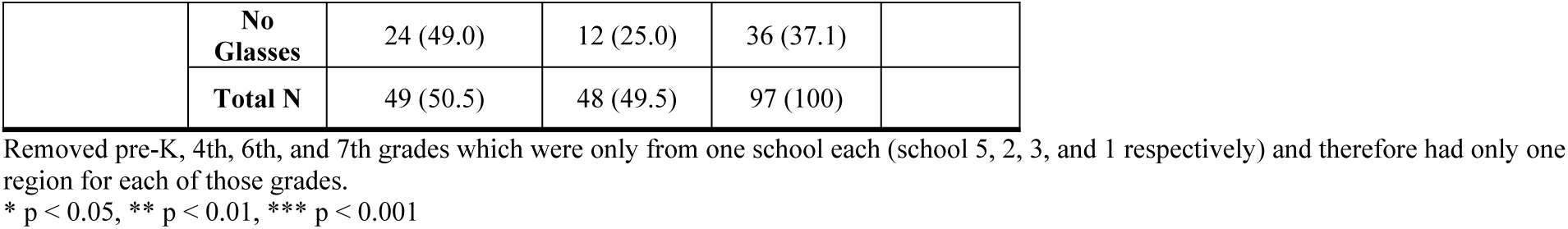
MobilEyes vision screening results by region.

The grade level analysis from Pre-K to 7^th^ grade students across all schools (irrespective of being from urban or suburban schools) demonstrated a significant difference between grades in failed screening proportion (p<0.001), ranging from 12.0% (Pre-K) to 43.2% (6^th^ grade). Similarly, the difference in proportion consented was significant (p = 0.021), ranging from 9.1% (Pre-K) to 44.5% (1^st^ grade). Difference in percentage of false positive screenings was not significant between grade levels (**S2 Table**). Controlling for potential bias by removing pre-K, 4th, 6th, and 7th grades resulted in similar significance in screening (p<0.001), consent (p = 0.022), and false positives (p = 0.319) **(Table 3)**.

**Table 3.** MobilEyes vision screening results across grade levels.

| Grade Level | Vision Screening (%) |  |  |  | Consent Process (%) |  |  |  | Glasses Prescription (%) |  |  |  |
| --- | --- | --- | --- | --- | --- | --- | --- | --- | --- | --- | --- | --- |
|  | Fail | Pass | Total | p-value | No Consent | Consent | Total | p-value | Glasses | No Glasses | Total | p-value |
| K | 36 (13.4) | 233 (86.6) | 269 (23.7) | <0.001*** | 22 (61.1) | 14 (38.9) | 36 (11.3) | 0.022* | 7 (50.0) | 7 (50.0) | 14 (14.4) | 0.319 |
| 1st | 119 (32.8) | 244 (67.2) | 363 (32.0) |  | 66 (55.5) | 53 (44.5) | 119 (37.4) |  | 31 (59.6) | 21 (40.4) | 52 (53.6) |  |
| 2nd | 31 (29.5) | 74 (70.5) | 105 (9.3) |  | 24 (77.4) | 7 (22.6) | 31 (9.7) |  | 5 (100.0) | 0 (0.0) | 5 (5.2) |  |
| 3rd | 75 (35.0) | 139 (65.0) | 214 (18.9) |  | 56 (74.7) | 19 (25.3) | 75 (23.6) |  | 13 (72.2) | 5 (27.8) | 18 (18.6) |  |
| 5th | 57 (31.1) | 126 (68.9) | 183 (16.1) |  | 41 (71.9) | 16 (28.1) | 57 (17.9) |  | 5 (62.5) | 3 (37.5) | 8 (8.2) |  |
| Total N | 318 (28.0) | 816 (72.0) | 1134 (100) |  | 209 (65.7) | 109 (34.3) | 318 (100) |  | 61 (62.9) | 36 (37.1) | 97 (100) |  |

Removed pre-K, 4th, 6th, and 7th grades which were only from one school each (school 5, 2, 3, and 1 respectively) and therefore had only one region for each of those grades.
\* p < 0.05, \*\* p < 0.01, \*\*\* p < 0.001

Among the 108 students total seen for a full eye exam, 81 (75.0%) were seen for routine annual follow-up and 41 of these (50.6%) were prescribed glasses. There were 27 (25%) students who needed earlier follow-up for a diagnosed ocular problem other than refractive error and all 27 of these students were prescribed glasses. There was no difference between urban and suburban schools in terms of sooner follow-up visit versus annual follow-up. There was a significant difference between the percentage of annual visit students requiring glasses (50.6%) and the earlier follow-up patients requiring glasses (100%) (p<0.001) (**Table 4**).

**Table 4.** MobilEyes vision screening results for follow-up.

| Needs follow-up earlier than 1 year | Glasses Prescription (%) |  |  |  |
| --- | --- | --- | --- | --- |
|  | Glasses | No Glasses | Total | p-value |
| No | 41 (50.6) | 40 (49.4) | 81 (75.0) | <0.001*** |
| Yes | 27 (100.0) | 0 (0.0) | 27 (25.0) |  |
| Total N | 68 (63.0) | 40 (37.0) | 108 (100) |  |

\* $p < 0.05$ , \*\* $p < 0.01$ , \*\*\* $p < 0.001$

Abnormalities diagnosed during the full eye exams included significant refractive error (hyperopia, myopia, astigmatism, anisometropia), amblyopia, optic disc cupping, exotropia, and convergence insufficiency.

## Discussion

In the current study, we aimed to examine the outcomes, challenges, and demographics of the MobilEyes program and compare these findings with similar school-based mobile eye care initiative programs in the United States. We found that 27.7% of all students failed vision screening, 32.3% of students that failed screening had parents return a consent form, and 37% of these that attended full eye exams were false positives. Ultimately 4.9% (N = 68) of the initial students screened (N = 1399) received glasses prescriptions. We found multiple statistically significant differences in screening across different schools (p < 0.001), different grades (p < 0.001), and between suburban and urban regions (p < 0.001). Difference in proportion consented was only statistically different across grade level (p = 0.021). Difference in proportion of glasses prescribed was only significant across region (p = 0.047) and students requiring follow-up earlier than 1 year vs 1 year follow up (p < 0.001).

To investigate the benefits of in-school “mobile” eye care for children that failed vision screening, Diao et al. compared this method to traditional referral to optometrists and pediatric ophthalmologists (18). They found that approximately 22,000 students fail vision screening in the Philadelphia Public School District each year, but only 23% are brought by parents/guardians to follow-up appointments with an optometrist or ophthalmologist. To improve this follow-up rate, they established a program that transported children to appointments by bus during school hours. The follow-up rate increased to 53% in the 2011-2012 school year. Then, as a final intervention, they formed a mobile eye care team, who provide follow-up appointments at school. This mobile team had the highest successful follow-up rate of 62%. The increase from a 53% to 62% follow-up rate was statistically significant (*P* = 0.036), offering strong evidence for the value and effectiveness of mobile eye care programs (18).

In order to establish an appropriate context in which to evaluate the MobilEyes program, we identified five mobile eye care programs with comparable methodologies. These programs were located in Florida (10), Virginia (5), Ohio (14), Baltimore (13), and Philadelphia (a more recent Philadelphia program than the one previously mentioned) (15). They generally adhered to the format of conducting vision screening and eye exams on-site at elementary, middle, and high schools. For students who did not pass the initial screening, programs performed full eye examinations for students either on the same day or later. When indicated, glasses were prescribed and dispensed, and referrals were made for further eye care. Consent forms were obtained at varying points in the process.

These programs have multiple variations in screening methods. Florida and Virginia programs used automatic photoscreening devices to determine refractive error (5, 10). The Ohio and Philadelphia programs used more tradition methods of screening such as visual acuity measurement via charts (i.e. Snellen, Allen), ocular alignment testing, and pen light inspection.(14, 15) The Baltimore program used both of these methods (13).

The three programs using automatic photoscreening devices in Virginia (5), Baltimore (13), and Florida (10) each had a similar initial vision screening failure percentage of 30%, 34%, and 32% respectively. The programs using traditional screening methods in Philadelphia(15) and Ohio (14) had lower screening failure percentages of 17% and 10% respectively. Philadelphia’s lower percentage, however, may be attributed to selection bias as “students were selected [for screening] by school nurses [only] if they had not been screened previously.”(15) Other programs screened all children present at school. A possible factor for the program in Ohio (14) is that the program uniquely had screening performed by optometrists or ophthalmic technicians rather than by school nurses or volunteers. Our screening with traditional methods also had a lower failure percentage of 27.7% compared to automatic photoscreening devices. Despite a similar method and screening team to Ohio, with a pediatric ophthalmologist, medical students, physician assistant students, and/or ophthalmic technicians, our percentage still appears to be higher.

These programs also differed on their timing of obtaining parental consent forms. The Baltimore, Ohio, and Florida programs obtained consent before screening, with screening percentages of 58%, 60%, and 72%.(10, 13, 14) Florida’s percentage was still higher despite including absent students within the no consent group (10). The Philadelphia program proceeded to the point of performing an eye exam and obtaining a glasses prescription but refrained from dispensing glasses until consent was obtained, resulting in a 77% consent percentage (15). In Virginia’s program, 2 of 3 districts employed a system where parents were sent a form to “opt-out” of potential eye examination and glasses prescription. Those students who did not return a signed opt-out form were assumed to be consented for participation in the program. The other district was a traditional “opt-in” consent form sent home after a failed screening. Virginia’s 73% consent percentage also included absent students within the no consent group (5). Our program screened students first and in cases of screening failure sent consent forms home with students for full exams. Our consent percentage of 32.3% was notably lower than the other studies. This may be due to our timing of consent selecting for vulnerable populations that were not already receiving eye care and failing exams or the method of “opt-in” consents that would need to make it home, be signed, and brought back to school.

Another point of variation between school-based vision screening programs was at the method of eye examination. Generally, optometrists performed the full eye examination for those children that did not pass initial screening. The Baltimore, Philadelphia, and Virgina programs’ full eye examinations consisted of nondilated fundus examination and noncycloplegic/manifest refraction (5, 13, 15). Baltimore and Virgina utilized slit lamp examinations as well (5, 13). The Florida program only specified that comprehensive vision exams were performed aboard the mobile vision clinic (10). The Ohio program administered 1% cyclopentolate and/or 1% tropicamide drops before cycloplegic retinoscopy and indirect ophthalmoscopy (14). Our study’s full eye examinations were completed by a pediatric ophthalmologist and also included cycloplegic refraction with cyclopentolate 1% ophthalmic drops with indirect ophthalmoscopy. Additionally, anterior segment exams were performed by pen light or slit lamp. The methodology of eye examinations may be a factor in the percent of students that were given glasses and consequently the false positive rate of programs.

Across programs, a false positive was defined as a failed vision screening that later underwent a full eye examination and did not result in a glasses prescription. The Virginia (5), Philadelphia (15), and Baltimore (13) programs had similar full eye examination methods, and of those vision screening failures that underwent a full eye examination, the percentage of students that needed glasses was similar—73%, 72%, and 72% respectively. This corresponds with vision screening false positivity percentages of 27%, 28%, and 28%. The programs in Ohio (14) and Florida (10) reported lower false positivity percentage of 16% and 15% respectively. Our study which had similar methodology Florida’s had a higher false positivity percentage of 37.0%. Our higher false positivity may be due to initial learning during the start of this program and prioritizing sensitivity over specificity. In the 2024-2025 school year we have started using the SPOT screener to determine better specificity. We have found this helpful in ruling out language barrier, age, and low-effort based limitations in screening.

## Strengths

One key strength of this study was analyzing demographic subgroups. This allowed us to see that the urban region had a statistically significant higher proportion of failed screening and lower proportion of false positivity compared to the suburban region. This was true both including and excluding grades with only one region. This may reflect the disparity between these populations resulting in less access to traditional health care and eye examinations, and increased number of students that have not yet been screened or given the prescription glasses that they need. The urban schools we visited had a greater percentage of socioeconomically disadvantaged students (94% in urban vs 61% - 73% in suburban schools) and minority students (95% - 98% in urban vs 54% - 66% in suburban schools) (17). It has been shown that lower income and minority youth are at greater risk for underdiagnosis and undertreatment of vision problems and 90% of visual impairment in 12 to 19 year olds was due to uncorrected refractive error (2, 5, 6). Therefore, there may be a greater need for mobile eye care programs in urban areas.

Another strength was our analysis by grade level which showed statistically significant differences in proportion of failed screening and consent. This was also consistent with both inclusion and exclusion of grades with students from only one school. However, these grades (PreK, 4^th^, 6^th^, and 7^th^) still have limited numbers in our sample and therefore limited conclusions we can draw from them. In NYS, screening is required in PreK, K, 1st, 3^rd^, 5^th^, and 7^th^. We found the screening failure percentage was highest at 6^th^ grade at 43%. However, 1^st^-5^th^ grades were similar at 29.5% to 35.0%, while PreK and K were lower at 12.0% and 13.4%. Consent percentages were highest in K-1^st^ and 7^th^ grade at 38.5% to 44.5% while lowest at 9.1% and 15.8% in PreK and 6^th^ grade. Percentages were similar across 2^nd^-5^th^ grades at 22.6% to 28.1%. Although 6^th^ grade had both the highest proportion of screening failure and one of the lowest proportion of consent, the number of 6th grade students in this analysis is too limited to determine if this is a possible area of need. This could be consistent with a California study showing myopia increasing from 6.1% in children 5 to <8 years old, to 31.2% in children 11 to <14 years old (19). Further complicating this issue, preteens often refuse to wear glasses due to concern of what peers may think (20, 21). Considering the increase in screening failure from PreK-K to 1^st^-5^th^ and the decrease in screening consent from K-1^st^ to 2^nd^-5^th^, an emphasis should be placed on continued screenings as children age, especially if future studies corroborate our 6^th^ grade findings. This analysis can guide future study in targeting areas of need by raising awareness at those specific grade levels with high screening failure and low consent.

The methodology of the program was also a strength with the screenings and full eye examinations (including cycloplegic refraction and indirect ophthalmoscopy) run by a team of ophthalmic technicians, volunteer medical students, and a supervising pediatric ophthalmologist. This was only done by one of the other five mobile eye care programs we compared with.

## Limitations

Although our study analyzed a few demographic subgroups, it did not directly cover race and ethnicity of individual students. We were able to indirectly comment on the disparities by comparing the total number of socioeconomically disadvantaged and minority students that attend the schools, however in the future the individuals that were screened, returned consent forms, and were prescribed glasses should be analyzed.

Despite the potential benefits of school-based vision screening programs, they face several challenges that can hinder their effectiveness. One of the most significant challenges is obtaining parental consent. One study found that using an opt-in consent method resulted in a 62% of children being screened compared to 89% with the opt-out method (22). Our opt-in method of consent may have limited the impact of our program. This same study found that in opt-in group only 8% opted out whereas 22% failed to return the consent form (22). This highlights that the primary limitation may be having the form signed and physically returned and not that parents are against their children being screened.

Studies have also shown that there are barriers that parents face that cause low follow-up rates. These often include lack of trust or understanding of the importance of visual findings on screenings or full eye exam visits. Additional barriers include transportation issues, unpredictable schedules, “living day to day” thinking only about basic needs, and questions about cost, insurance, or privacy (5, 23–25). These factors can lead to low consent rates, which in turn can limit the number of children who receive screening, full eye examinations glasses, and referral for care.

## Next steps

Further research is needed to refine future programs and aid the specific needs of students. Trials of the different methodologies for screening and full eye examinations would be helpful for lowering false positivity. Urban and suburban subgroups were analyzed based on students enrolled in those schools that are considered socioeconomically disadvantaged students and minority students. Analyzing the students screened that fall into these groups would allow for more accurate interpretation of urban and suburban subgroups. The reasoning for significant differences between grade levels should be further explored through questions on previous eye exams or screenings of these students and questionnaires for parents on reasons for consent decisions. Students should continue to be followed to understand the future impact of our program on follow-up visits and measurable outcomes such as changes in school performance and behavior.

In the future, we plan to improve the program in several areas via expansion and education. Efforts to improve the consent process would increase the number of students we can examine and subsequently the power of the data. Potential methods to improve this process include using an opt-out method and providing various modalities to distribute and receive consent, such as an electronic or verbal method (via the phone with a legal guardian). In addition to improving the consent process, we plan to expand our partnership with more schools in the Greater Rochester region with emphasis on low income populations in both urban and rural settings.

Providing patient-centered education for legal guardians may improve the follow-up rates after failed vision screening. The education would be a brief introduction to eyecare, the effect of poor vision on a child’s development, and the importance of following up for an eye exam. We believe these steps will enhance the impact and sustainability of our efforts to address visual impairment in grade school children.

## Conclusion

Our data highlights the increased disparities and need for school screening and full eye examinations in urban regions. The prevalence of visual impairment among these communities underscores the value of school-based mobile eye care programs due to its improved follow-up and increased glasses prescriptions and eye care to children in need. There is also a need to explore why different grade levels have different proportions of screening failure and consent to better address the needs of students. However, a consistent challenge encountered across programs including our own is variable false positivity percentages and low percentage of parental consent for in-school eye exams. Furthermore, the variability in screening methods, criteria, and follow-up procedures across different programs and states further highlights the need for more standardized and evidence-based approaches to pediatric vision screening.

Overall, effective communication and development of mutual trust are crucial at every stage of the process, from obtaining parental consent, to conveying screening results, addressing parental concerns for follow-up care. MobilEyes and other similar programs can continue to fine-tune their approach to achieve an essential service in under resourced settings that reduces visual impairment and promotes optimal health and development among children.

## Data Availability

All relevant data are within the manuscript and its Supporting Information files. Individual student data cannot be given due to FERPA, only aggregated data is included here.

## Acknowledgements

The authors would like to thank Research to Prevent Blindness and the Mother Cabrini Health Foundation for their support. Additionally, the authors express their gratitude to the six schools and the nurse at each site that participated in the MobilEyes program during the 2022-2023 academic year.

## Supporting Information

**S1 Table. Screening results by region without confounder adjustment.** * p < 0.05, ** p < 0.01, *** p < 0.001

**S2 Table. Screening results across grade levels without confounder adjustment.** * p < 0.05, ** p < 0.01, *** p < 0.001

**S3 Table. MobilEyes vision screening results for follow-up by region.** * p < 0.05, ** p < 0.01, *** p < 0.001

